# Study Protocol and Rationale of “The UP Project”: Evaluating the Effectiveness of Active Breaks on Health Indicators in Desk-Based Workers

**DOI:** 10.1101/2023.12.28.23300545

**Authors:** Carlos Cristi-Montero, Ricardo Martínez-Flores, Juan Pablo Espinoza-Puelles, Laura Favero-Ramirez, Natalia Zurita-Corvalan, Ignacio Castillo Cañete, Jaime Leppe, Gerson Ferrari, Kabir Sadarangani, Jorge Cancino-López, Sam Hernandez-Jaña, Tuillang Yuing Farias, Vanilson Batista Lemes, Fernando Rodríguez-Rodríguez, Caroline Brand

**Author notes:** All three researchers contributed equally to this study (Carlos Cristi-Montero, Ricardo Martínez-Flores, Caroline Brand). **Corresponding** Ph.D. Carlos Cristi-Montero, IRyS Group, Physical Education School, Pontificia Universidad Católica de Valparaíso, Valparaíso, Chile.

## Abstract

**Background:** Excessive sedentary time has been negatively associated with several health outcomes, and physical activity alone does not seem to fully counteract these consequences. This panorama emphasizes the essential of sedentary time interruption programs. “The Up Project” seeks to assess the effectiveness of two interventions, one incorporating active breaks led by a professional and the other utilizing a computer application (self-led), of both equivalent duration and intensity. These interventions will be compared with a control group to evaluate their impact on physical activity levels, sedentary time, stress perception, occupational pain, and cardiometabolic risk factors among office workers.

**Methods:** This quasi-experimental study includes 60 desk-based workers from universities and educational institutes in Valparaiso, Chile, assigned to three groups: a) booster breaks led by professionals, b) computer prompts that are unled, and c) a control group. The intervention protocol for both experimental groups will last 12 weeks (only weekdays). The following measurements will be performed at baseline and post-intervention: cardiometabolic risk based on body composition (fat mass, fat-free mass, and bone mass evaluated by DXA), waist circumference, blood pressure, and resting heart rate. Physical activity and sedentary time will be self-reported and objectively assessed using accelerometry. Questionnaires will be used to determine the perception of stress and occupational pain.

**Discussion:** Governments worldwide are addressing health issues associated with sedentary behavior, particularly concerning individuals highly exposed to it, such as desk-based workers. Despite implementing certain strategies, there remains a noticeable gap in comprehensive research comparing diverse protocols. For instance, studies that contrast the outcomes of interventions led by professionals with those prompted by computers are scarce. This ongoing project is expected to contribute to evidence-based interventions targeting reduced perceived stress levels and enhancing desk-based employees’ mental and physical well-being. The implications of these findings could have the capacity to lay the groundwork for future public health initiatives and government-funded.

## Background

Sedentary behavior is defined as any waking behavior with an energy expenditure of ≤1.5 METs while sitting or lying down (1). This behavior has been negatively associated with various health markers, such as, sarcopenia, osteopenia, diabetes, hypertension, metabolic syndrome, cardiovascular disease, overweight, obesity, and stress (2–7). This scenario is even worse in desk-based workers exposed daily to a high amount of sedentary time, increasing the frequency and duration of work disability, which implies unfavorable implications for the worker, company, and society (8). Consequently, interrupting sedentary time with physical activity emerges as a valuable strategy for addressing sedentary behavior (9–11).

In this sense, advocating for workplace programs that promote physical activity and reduce the prolonged time spent in sedentary activities becomes a practical proposition in occupational health (involving both physical and mental aspects of well-being). The World Health Organization characterizes occupational health as a highly interdisciplinary effort to safeguard and enhance workers’ health by preventing and managing work-related diseases and accidents (12). A key point to emphasize is that scientific evidence underscores how certain detrimental health outcomes associated with sedentary behaviors seem to be independent of individuals’ physical activity levels. Thus, interventions based on physical activity alone would not be entirely effective in counteracting the adverse effects of sedentary behavior on health (9). In this regard, strategies for reducing sedentary time have been associated with decreased body fat, increased fat-free mass, decreased abdominal fat, resting heart rate, and blood pressure (10,13–15).

Given the potential hazards associated with prolonged sitting at work and physical inactivity, including increased stress and occupational pain (16–18), it is imperative to examine workplace interventions that can mitigate these behaviors and promote overall health and well-being (19). The literature has mainly proposed two types of active breaks to interrupt sedentary behavior in desk-based workers: “Booster Breaks” (B-B) and “Computer Prompts” (C-P) (20–22). On the one hand, B-B can be defined as guided, programmed, and performed active breaks in the workplace, with an average duration around of 15 minutes (20). This type of break benefits individuals’ physical and mental health by reducing stress, improving social interactions at work, increasing physical activity levels, and decreasing body mass index (BMI) (20,21). On the other hand, C-P is characterized by unguided breaks performed through the software on the work computer, with a short duration (e.g., 2-3 minutes), accumulating around 15 minutes during the whole working day (20). Two systematic reviews with meta-analyses have concluded that this type of break can reduce sedentary behavior and increase physical activity levels, but the body of evidence comparing B-B and C-P is still limited (22,23).

Thereby, the current project pretends to fill gaps in the existing literature on this topic in an under-researched geographical world region. First, previous studies investigating B-B and C-P protocol parameters used physical activity questionnaires, pedometers, BMI, and waist circumference measures to analyze body composition (20,22). However, to date, scarce evidence has evaluated movement behavior by accelerometry and dual-energy X-ray absorptiometry (DXA) to accurately measure participants’ body composition using the same protocol. Thus, we will combine traditional and objective measurements to evaluate our outcomes.

Second, this study introduces an innovative approach by contrasting two types of interventions (B-B and C-P), which will be equivalent in duration and intensity. However, these interventions differ in their structures. B-B will be conducted continuously for 14-16 minutes, while C-P will consist of brief 2-minute breaks scattered throughout the 8-hours workday (averaging 14-16 minutes). By assessing the differentiated effects of these breaks, we aimed to identify effective strategies to improve the health of office workers.

Third, previous scientific evidence has shown that diverse variables can affect our main outcomes, encompassing factors such as sleep time and quality, socioeconomic status, smoking, and eating habits (24–26). Consequently, it is imperative to explore these interrelated factors using a more comprehensive approach (27). Initially, these variables will be used as covariates to explore their influence on study outcomes.

Finally, it is crucial to take public health measures to contribute to the reduction of sedentary behaviors in workers. Thus, we anticipate that our findings could contribute to the formulation of evidence-based interventions that can be deployed or not to improve the health and well-being of this population.

Therefore, this manuscript aims to describe the design and methods of “The Up Project,” which seeks to establish the efficacy and differences between two interventions addressed for interrupting sedentary time in desk-based workers. The principal outcomes will include variations in cardiometabolic risk factors, sedentary and physical activity levels, perception of stress, and occupational pain. Accordingly, our primary hypothesis is that both intervention groups will yield better results compared to the control group. However, we expect no discernible differences between the intervention groups due to their equivalence in duration and intensity.

## Methods

### Study design and ethical considerations

“The Up Project” comprises a quasi-experimental study in which the sample was selected through cluster sampling, with workplaces serving as the basis for establishing both interventions and control groups (ClinicalTrials.gov identifier: NCT05844267). This project has been approved by the Ethics Committee of Pontificia Universidad Católica de Valparaíso (BIOEPUCV-HB 580–2023) and will be conducted following the Declaration of Helsinki and the guidelines of the SPIRITS (Standard Protocol items for intervention trials) checklist (28). Additional information is provided in the supplementary material. Written consent will be obtained from the participants before beginning and will be guarded and stored by the principal investigator to maintain the confidentiality of the participants. All protocol modifications will be communicated and registered on ClinicalTrials.gov.

### Training of personnel and quality control

Our research group will organize training sessions for investigators and key personnel prior to data collection. Quality control and rigorous standardization of measurement protocols across centers are critical for the success of intervention studies. All investigators from our research group shared responsibility for quality control. The principal investigators will oversee regulatory compliance, protocol adherence, data accuracy, personnel training, and regulatory document management. During data entry, key variables will be checked for accuracy with assigned interval controls. A review will be required for any data entered outside the preset ranges. The principal investigators make the final decision to terminate the trial.

### Sample recruitment

Desk-based workers (60 in total) will be recruited voluntarily from universities and educational institutes in the Valparaiso region, Chile. We will extend an open invitation to all desk-based workers of the Pontificia Universidad Católica de Valparaíso (PUCV) in the Faculty of Philosophy and Education and the central office located in the Central Campus. The control group sample will be obtained from Naval Academy’s desk-based workers. It is important to note that the participants of the control group are civilians or retired marines who do not undergo any type of military training or physical tests. Subsequently, our research team will be visiting their offices to conduct face-to-face recruitment. During this process, the participants will be provided a detailed description of the scientific background, research objectives, and safety measures by our research group. We aim to ensure that all participants clearly understand our research and feel comfortable participating in our study. Once accepted, they will be assigned to a group according to the work site. Upon completion of the study, the participants will be given a full report on changes in body composition, physical activity levels, and sedentary time. After the data collection period, the control group will have the opportunity to participate in the same intervention program.

#### Inclusion and exclusion criteria

To be included in this study, participants had to be desk-based women and men workers, between 18 and 60 years of age, who belong to an educational institution in the Valparaiso region, with a full-time contract.

As exclusion criteria will be considered participants who do not have a full-time job, individuals with physical limitations for physical exercise or undergoing weight loss treatment, pregnant and lactating (in the first 6 months postpartum) women (29), or pacemaker users (30). In addition, participants with a participation frequency lower than 70% in the protocol will not be considered; however, a sensitivity analysis will be performed based on the intention-to-treat principle.

#### Sample size calculation

The sample size was calculated using the G*Power software. This calculation considered the repeated measures of factorial analysis of variance (ANOVA), with an effect size (f) of 0.20, alpha value of 0.05, and beta of 0.80. The study involved three groups and two evaluation time points (pre/post), with an estimated follow-up loss of 10%. Consequently, 60 participants will be distributed as 20 participants per group: two experimental groups (B-B led by professionals and unled C-P) and one control group. The groups will be sex balanced because of the predominance of women in this type of work. In this sense, for selecting participants, we used the sex ratio of workers in group B-B (the lowest men proportion). In this site, there is a proportion of 27.1% of men and 72.9% of women; thus, our sample will try to approach this proportion in the two remaining groups.

#### Procedures

The project will be conducted in three stages, comprising two visits to the Physical Performance and Health Laboratory of the PUCV (stage one and stage three) and one visit to the participants’ offices (stage two). The first stage will consist of a pre-intervention measurement session in our laboratory, in which cardiometabolic risk factors, stress perception, occupational pain, physical activity, sedentary time at work, 24-hours behavior, eating and smoking habits, and sociodemographic information will be evaluated. In addition, participants will be given an accelerometer, which will be removed seven days later, and the intervention will begin once the accelerometer is removed. In the second stage (sixth week of intervention), the three groups will receive accelerometers to re-evaluate their daily physical activity intensity, sedentary time, and breaks in sedentary time. Finally, the third stage (post-intervention measurement) will consist of a second visit to our laboratory, where all variables will be re-evaluated. The anonymized and raw data obtained will be made available by the corresponding author upon reasonable request. A diagram of the study design, sample, and measures is presented in Figure 1.

**Figure 1.**
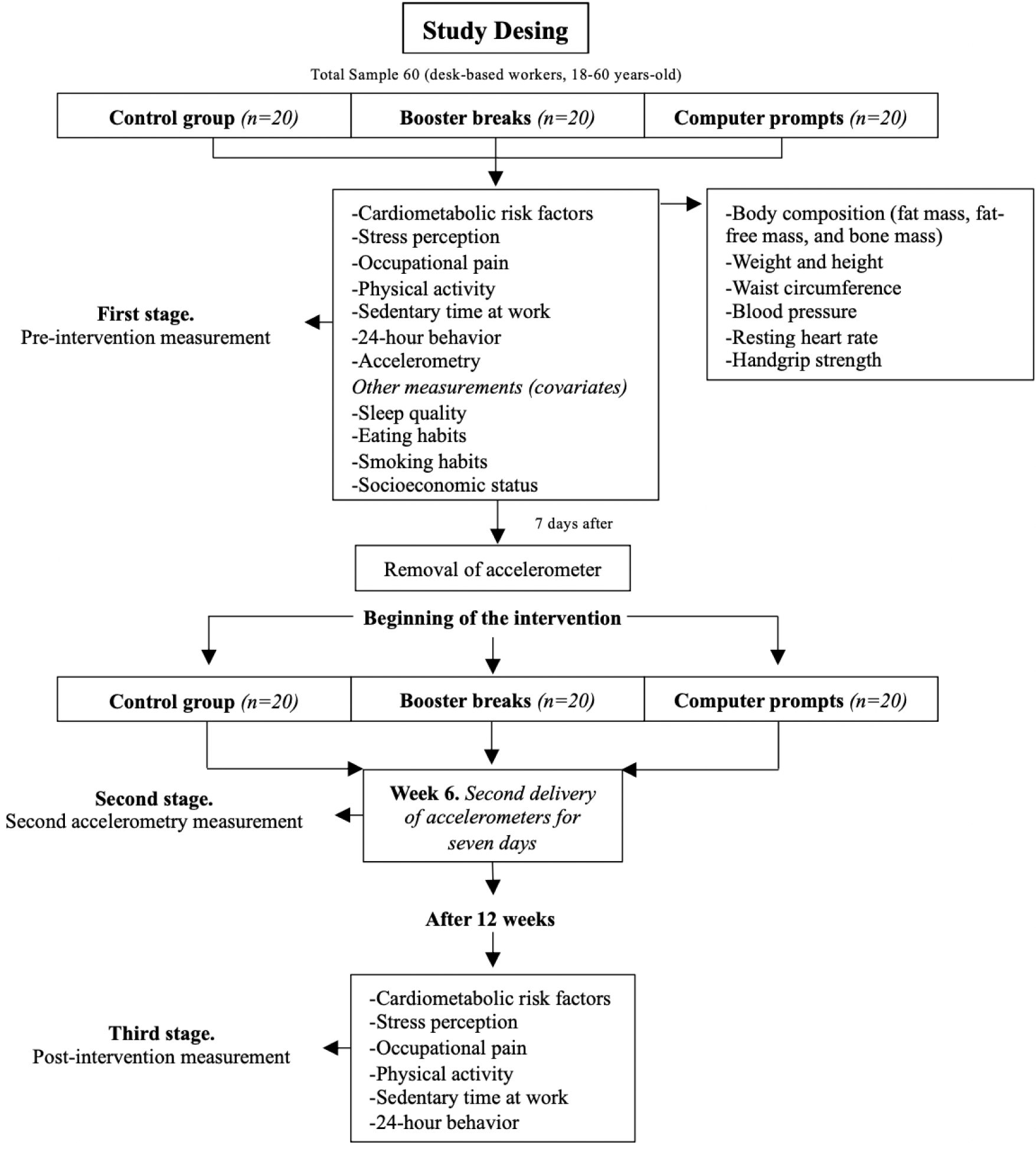
Study Design.

### Main variables

In the first stage, upon arrival at the laboratory, participants will be asked to read and sign an informed consent form and then begin the measurements. First, the participant will be asked to complete questionnaires (stress perception, occupational pain, physical activity, sedentary time at work, 24-hours behavior, eating habits, smoking habits, and socioeconomic status). Subsequently, the participants will be asked to enter the DXA box in the laboratory, where their height, weight, and waist circumference will be assessed. Once these measurements have been taken, the participant will be asked to lie on the scanner table for analysis. Upon completion of the scanner (estimated duration of 7 minutes), the subject’s resting heart rate will be determined while they remain in a lying down position. Thereafter, the individual will be requested to sit down, and their blood pressure will be measured in their left arm twice consecutively. Lastly, the accelerometer will be provided to the participant for a period of seven days, for the initial measurement.

In the second stage, our research team members will go to the participants’ workplaces for the second delivery of accelerometers for seven days. In the third stage, questionnaires will be applied to the participants (stress perception, occupational pain, physical activity, and sedentary time at work). Subsequently, measurements of cardiometabolic risk factors will be made with the previously mentioned protocol. All measurements will be carried out between 16:00 and 20:00 for the convenience of the participants due to their work schedule.

#### Cardiometabolic Risk Factors

DXA scans (General Electrics, model Lunar, series 212069) will be used to assess body composition (fat mass, fat-free mass, and bone mass). We will use a digital balance (OMRON HN-289-LA) and a portable stadiometer (SECA, model 213, GmbH) to measure weight and height. After exhaling, waist circumference will be measured using a Lufkin metallic tape measure (W606PM). Additionally, we will use a digital monitor (model HEM-7120, OMRON) to measure blood pressure and a pulse oximeter (model Prince-100B5, Heal Force) to determine the resting heart rate. Finally, we will measure handgrip strength using a digital dynamometer (JAMAR Plus Digital Hand Dynamometer). This will be evaluated two times per hand, with the person standing and the arm extended parallel to the body. Previous studies indicate that these measurements are related to the relative risk of cardiometabolic factors (24–26,31).

#### Physical Activity, Sedentary Time, and Sleep Time

An ActiGraph GT3X + accelerometer (ActiGraph, Illinois, USA) will be used to objectively monitor physical activity, sedentary behavior, and sleep time. The accelerometer has been validated for assessing physical activity, sedentary behavior, and sleep quality (32,33).

Participants will be encouraged to wear the accelerometer 24 hours a day for 7 days. The minimum amount of accelerometer data that will be considered acceptable will be 600 min of valid daily monitor wear on at least 4 days (34,35). After the last day of data collection, the accelerometers will be returned to the research team for processing using the GGIR package in RStudio (36).

To complement the accelerometer data, we will administer four questionnaires: a) the Physical Activity Questionnaire (GPAQ), a standardized tool recognized by the World Health Organization (WHO) that provides valuable information on physical activity levels globally (37), b) a single physical activity question to identify the level of physical activity through a single item (38), c) the Occupational Sitting and Physical Activity Questionnaire (OSPAQ) will be used to determine the proportion of time participants spend on sedentary behavior during work hours (39), and finally, d) the Daily Activities Behavior Questionnaire (DABQ) to gather additional information on participants’ physical activity, sedentary behavior, and sleep habits over 24-hours (40). Questionnaires b, c, and d will be validated in Spanish in a parallel study by our investigation group. The combination of objective measurement tools and standardized questionnaires will enable an accurate and comprehensive assessment of physical activity levels, sedentary behavior, and sleep quality. All of these variables will allow us to analyze the effect of reducing sedentary time and its impact on the study’s objective (41,42). The data collected will be crucial for analyzing the results and effectiveness of interventions among office workers.

#### Stress Perception

The Perceived Stress Scale Questionnaire (PSS-14) will measure stress perception in various situations. This questionnaire was initially validated by Cohen, Kamarck, and Mermelstein in 1983 (43) and has been adapted for use in Chile by Erik Marín (44). The PSS-14 questionnaire assesses an individual’s perception of different situations in which their stress levels may be influenced. The questions focused on the last month and are rated on a Likert scale ranging from 0 (never) to 4 (very often).

#### Occupational pain

The Nordic Musculoskeletal Questionnaire, a standardized tool recommended by the Institute of Public Health of the Chilean Ministry of Health, will measure occupational pain. This questionnaire is specifically designed to detect musculoskeletal symptoms related to desk-based work, including pain in the neck, shoulder, thoracic spine, wrist, lumbar spine, hip, knee, and ankle (45). With the implementation of this tool, we will be able to assess both the general and specific pain experienced by participants in the workplace. This method provides a comprehensive evaluation of the prevalence and severity of musculoskeletal pain, enabling a thorough analysis of the overall impact on employees’ well-being (17,46,47).

#### Other measurements (covariates)

Previous studies have established that cardiometabolic risk factors are not only affected by body composition, physical activity, or sedentary behavior but can also be influenced by various healthy lifestyle habits (24–26). Therefore, we identified a series of related factors that will be included as covariates in our statistical analyses to assess their impact on our outcomes.

We will use an ActiGraph accelerometer to measure sleep quality (32,33). This variable plays a fundamental role in cardiovascular health (48). Lack of quality sleep can increase blood pressure and inflammation by increasing cortisol levels and decreasing immunity, elevating the risk factors for cardiovascular problems (48).

The questionnaire on adherence to the Mediterranean diet will be applied to assess eating habits, specifically designed to measure participants’ adherence to it and determine its relationship with cardiometabolic factors (49,50).

The Fargerstörm test, which scores various aspects of smoking habits on a Likert scale and enables us to determine the level of dependence on this substance, will be used to assess smoking habits (51).

Furthermore, we will obtain information on the participants’ socioeconomic status using a questionnaire on the distribution and average autonomous income of households by income deciles. This questionnaire provides essential data on the per capita income of the participants and follows the guidelines of the “Asesoría Técnica Parlamentaria” of the Chilean government (52). These measures are considered relevant as predictive cardiometabolic risk factors (50,53,54). In addition, sociodemographic questions will be asked to characterize the study sample.

### Protocol interventions

#### Computer prompts group (unled breaks)

Active breaks during the workday will be guided by an application called “Ponte de Pie por tu Salud” developed by the Chilean Ministry of Health (https://unchilemassano.minsal.gob.cl/#). The exercises included in these breaks focus on stretching, mobility, and muscular resistance with light to moderate intensity (i.e., Rate of Perceived Exertion [RPE] 3-4) following the Chilean Safety Association (*Asociación Chilena de Seguridad* - ACHS) recommendations (55,56). An example of the exercises to be performed can be found in the following link (https://unchilemassano.minsal.gob.cl). The sessions will begin with a warm-up and end with a cool-down, following the ACHS guidelines. The application will be configured to generate 2-minute breaks every hour for 8 hours per day, ensuring that participants accumulate an average of 14-16 minutes of active breaks each day. By implementing this approach, we can ensure that both interventions are distributed equally over time.

#### Booster Breaks group (led breaks)

We will implement active breaks lasting approximately 14 to 16 minutes every working day for 12 consecutive weeks, for five workdays, excluding weekends. These breaks will consist of breathing exercises, stretching exercises, and exercises recommended by the ACHS (55,56). They will be similar in intensity to exercises proposed in the “Ponte de Pie por tu Salud” application. Each session will be divided into three parts: warm-up (2 minutes), main work (12 minutes), and cool down (1 minute). The intensity of the exercises will range from light to moderate (RPE 3-4), and at the end of each session, we will evaluate intensity perception using Borg’s scale (57). Trained personnel from our research team will conduct all the sessions throughout the intervention.

During the warm-up period, we will include breathing and muscle activation exercises that will last approximately 2 minutes. For the main work, we will target four main muscle groups: i) neck and shoulders, ii) arms and wrists, iii) lumbar back and hips, and iv) knees and ankles, which will be distributed and combined according to Table 1.

**Table 1.** Graphical example of the time distribution of muscle groups in one month of intervention.

| Days | Week 1 | Week 2 | Week 3 | Week 4 |
| --- | --- | --- | --- | --- |
| Monday | i 70% - B 30% | ii 70% - A 30% | i 70% - B 30% | ii 70% - A 30% |
| Tuesday | iii 70% - D 30% | iv 70% - C 30% | iii 70% - D 30% | iv 70% - C 30% |
| Wednesday | i 70% - C 30% | iii 70% - A 30% | i 70% - C 30% | iii 70% - A 30% |
| Thursday | ii 30% - D 70% | iv 30% - B 70% | ii 30% - D 70% | iv 30% - B 70% |
| Friday | iv 30% - A 70% | i 30% - D 70% | iv 30% - A 70% | i 30% - D 70% |

The program will be cycled over the next two months. For more information on the sequence of exercises, please contact the corresponding author.

To prevent participant dropout and maintain motivation throughout the intervention, both variations and combinations of the main muscle groups to be worked on will be introduced. The two muscle groups will be combined daily, with 70% of the time allocated to the first group and 30% to the second group. The order and time percentages will be reversed the following week. In the second month, elastic bands and canes will be incorporated to support the exercise performance. The main work session will last nearly 12 minutes.

It is important to note that although B-B includes exercises that differ from those of the application, the intensity will be the same. Borǵs scale will be employed for this purpose to ensure comparability between the interventions. In this sense, our investigation group conducted a pilot test of the selected exercises to determine the correct intensity in relation to the exercises proposed by the application. In addition, the staff in charge of conducting the group B-B active breaks ensured that the sessions were performed between light and moderate intensity (i.e., RPE 3-4).

#### Control group

The control group will not have any type of intervention; they will be asked to continue with their normal life habits. At the end of the study, participants will be invited to participate in a physical activity program at their workplace.

#### Follow up of participants

For the C-P group, the computer application will record the number of times the breaks will be accepted, rejected, or postponed, and it will generate a weekly summary. Furthermore, a member of our research group will consistently make phone calls to ensure that everything remains in order. In the case of the B-B group, all sessions will be recorded, with attendance tracking for all participants. Meanwhile, for the control group, a research group member will periodically remind them of the recommendations provided at the beginning of the 12 weeks.

In case of desertion for any reason (i.e., leaving the country, change of job, impossibility of measurements), a sensitivity analysis will be performed with and without intention to treat. In case of adverse effects (i.e., injuries or health problems), the situation will be evaluated, and the necessary decisions will be taken.

### Data analysis plan

Owing to the characteristics of the study (i.e., cluster for conformation of the groups), allocation concealment was not possible. However, the blinding of the evaluator is detailed in the study records (more information in ClinicalTrials.gov, identifier: NCT05844267).

Descriptive data from participants will be summarized and presented as means and standard deviations for continuous variables, and frequencies and percentages for categorical variables. Appropriate statistical tests, such as T-tests or Chi-square tests, will be used to determine differences in baseline characteristics and sex differences where applicable. The normality of the variables will be explored using the Shapiro-Wilk test, and visual analysis with Q-Q plots and distribution of residuals will be applied. If the criteria associated with missing at random data are met, the relevance of the imputation will be evaluated.

Analysis of variance (ANOVA) or analysis of covariance (ANCOVA) will be applied depending on the objectives of the study. The inter-individual response of the participants will also be analyzed if deemed necessary. Possible mediating and moderating effects will be explored. Any additional analysis that the researchers consider necessary will be sought.

In parallel, sensitivity analyses will be performed according to the intention-to-treat principle. All analysis models will also adjust for relevant covariates such as sex, age, socioeconomic status, eating habits, smoking habits among others depending on their relevance in the model studied.

## Discussion

### Expected results and transfer to the work context

Reducing health problems associated with sedentary behavior is a crucial concern for governments worldwide (60). While several strategies are emerging to mitigate excessive sedentary time in desk-based workers, comprehensive research examining multiple outcomes and protocols is lacking (22,23,27). In this context, our study will compare two intervention types (B-B and C-P) that are equivalent in duration and intensity but differ in distribution. This design, which is more realistic for future interventions, will allow us to compare a) the effectiveness of a continuous break protocol versus an interval-based one and b) the effectiveness of professional-directed pauses versus autonomous breaks directed by a computational application.

Furthermore, this research helps a gap in the literature by concentrating on three pivotal facets of participants’ health. Firstly, we hope that employing DXA for body composition measurement and accelerometry for assessing sedentary and physical activity time could enhance methodological precision in this field. Secondly, we aim to explore effective strategies for enhancing cardiometabolic risk markers in desk-based workers through a more comprehensive and innovative approach. Thirdly, our objective is to contribute to developing evidence-based interventions aimed at diminishing perceived stress levels and occupational pain experienced by workers. Addressing these three indicators could be significant for companies and public health policies. In this line, our hypothesis is that both intervention groups will yield better results compared to the control group.

However, this trial has limitations. First, randomization will not be possible due to the intervention’s characteristics as well as blinded allocation. Second, stress perceived by desk-based workers will be measured through a questionnaire, and exercise intensity will be gauged by RPE. In both instances, it would be desirable to incorporate complementary physiological indicators, such as cortisol levels and heart rate, respectively. Additionally, our assessment of cardiometabolic risk factors does not currently encompass a lipid profile.

This study also has strengths to highlight. First, although the OSPAQ, DABQ, and single item of the physical activity questionnaire are not validated in Spanish language, our research group is actively working on these validations in a parallel study. Second, the intervention during 12 weeks already allows to observe behavioral change (61), which could extend in time the effects of the interventions carried out after the study’s end. Thirdly, integrating both objective and traditional measures for cardiometabolic risk, physical activity, and sedentary lifestyle will enable the creation of a more comprehensive model. This model aims to enhance our understanding of how interventions impact these indicators collectively. In this regard, it is important to remark on the use of DXA for body composition measurements. Fourth, the determination of specific covariates that may affect our outcomes in important ways. These covariates will allow us to adjust our analysis model in order to comprehensively explore the effects of the intervention proposed in this study.

The results of this project can serve as a basis for future interventions, and government and company programs aimed at improving the health of desk-based workers. Promoting these strategies is essential, given that prolonged sitting in the workplace combined with physical inactivity is a crucial risk factor for workers’ physical, metabolic, and mental health.

In summary, “The UP Project” aims to identify efficient ways to improve markers of cardiometabolic risk, perceived stress and occupational pain in office workers through breaks aimed at interrupting sedentary time during the workday. We hope that this effort will mark a breakthrough in the search for protocols to improve the health of office workers and contribute with scientific evidence that will contribute to the development of public policies to combat sedentary time.

## Authors’ contributions

CCM and CB took the lead in designing the study. RM-F, CCM, and CB prepared the original manuscript. JPEP, LF-R, NZ-C, ICC, JL, VBL, GF, KS, JC-L, SH-J, TYF, and F, R-R provided expert input and support overall for the writing of this manuscript with particular emphasis on design. All authors read and approved the final manuscript.

## Availability of data and materials

Not applicable.

## Ethics approval and consent

To participate Ethics Committee of Pontificia Universidad Católica de Valparaíso has provided ethical approval to conduct this study (BIOEPUCV-HB 580–2023). All participants provided informed written consent to participate in the study.

## Supporting information

Supplementary Material SPIRIT Checklist

## Data Availability

All data produced in the present study are available upon reasonable request to the corresponding author

## References

1. Tremblay MS, Aubert S, Barnes JD, Saunders TJ, Carson V, Latimer-Cheung AE, et al. Sedentary Behavior Research Network (SBRN) – Terminology Consensus Project process and outcome. Int J Behav Nutr Phys Act. 2017 Jun 10;14(1):75.

2. Chauntry AJ, Bishop NC, Hamer M, Kingsnorth AP, Chen YL, Paine NJ. Sedentary behaviour is associated with heightened cardiovascular, inflammatory and cortisol reactivity to acute psychological stress. Psychoneuroendocrinology. 2022 Jul;141:105756.

3. Koedijk JB, van Rijswijk J, Oranje WA, van den Bergh JP, Bours SP, Savelberg HH, et al. Sedentary behaviour and bone health in children, adolescents and young adults: a systematic review. Osteoporos Int. 2017;28(9):2507–19.

4. NCD Risk Factor Collaboration (NCD-RisC). Heterogeneous contributions of change in population distribution of body mass index to change in obesity and underweight. eLife. 2021 Mar 9;10:e60060.

5. Olsen MH, Angell SY, Asma S, Boutouyrie P, Burger D, Chirinos JA, et al. A call to action and a lifecourse strategy to address the global burden of raised blood pressure on current and future generations: the Lancet Commission on hypertension. Lancet Lond Engl. 2016 Nov 26;388(10060):2665–712.

6. Patterson R, McNamara E, Tainio M, de Sá TH, Smith AD, Sharp SJ, et al. Sedentary behaviour and risk of all-cause, cardiovascular and cancer mortality, and incident type 2 diabetes: a systematic review and dose response meta-analysis. Eur J Epidemiol. 2018 Sep;33(9):811–29.

7. Sánchez-Sánchez JL, Mañas A, García-García FJ, Ara I, Carnicero JA, Walter S, et al. Sedentary behaviour, physical activity, and sarcopenia among older adults in the TSHA: isotemporal substitution model. J Cachexia Sarcopenia Muscle. 2019 Feb;10(1):188–98.

8. Bailey DP. Sedentary behaviour in the workplace: prevalence, health implications and interventions. Br Med Bull. 2021 Mar 25;137(1):42–50.

9. González K, Fuentes J, Márquez JL. Physical Inactivity, Sedentary Behavior and Chronic Diseases. Korean J Fam Med. 2017 May;38(3):111–5.

10. de Rezende LFM, Rodrigues Lopes M, Rey-López JP, Matsudo VKR, Luiz O do C. Sedentary behavior and health outcomes: an overview of systematic reviews. PloS One. 2014;9(8):e105620.

11. Ekelund U, Steene-Johannessen J, Brown WJ, Fagerland MW, Owen N, Powell KE, et al. Does physical activity attenuate, or even eliminate, the detrimental association of sitting time with mortality? A harmonised meta-analysis of data from more than 1 million men and women. The Lancet. 2016 Sep 24;388(10051):1302–10.

12. World Health Organization. Sixtieth World Health Assembly. Workers health: global plan of action. 2007.

13. Alansare AB, Bates LC, Stoner L, Kline CE, Nagle E, Jennings JR, et al. Associations of Sedentary Time with Heart Rate and Heart Rate Variability in Adults: A Systematic Review and Meta-Analysis of Observational Studies. Int J Environ Res Public Health. 2021 Aug 12;18(16):8508.

14. González-Gross M, Meléndez A. Sedentarism, active lifestyle and sport: Impact on health and obesity prevention. Nutr Hosp. 2013 Sep;28 Suppl 5:89–98.

15. Larsen BA, Allison MA, Kang E, Saad S, Laughlin GA, Araneta MRG, et al. Associations of physical activity and sedentary behavior with regional fat deposition. Med Sci Sports Exerc. 2014 Mar;46(3):520–8.

16. Dėdelė A, Miškinytė A, Andrušaitytė S, Bartkutė Ž. Perceived Stress among Different Occupational Groups and the Interaction with Sedentary Behaviour. Int J Environ Res Public Health. 2019 Nov 20;16(23):4595.

17. Gilson ND, Burton NW, van Uffelen JGZ, Brown WJ. Occupational sitting time: employees’ perceptions of health risks and intervention strategies. Health Promot J Aust Off J Aust Assoc Health Promot Prof. 2011 Apr;22(1):38–43.

18. D J, M Z, V J, S O. Physical risk factors for developing non-specific neck pain in office workers: a systematic review and meta-analysis. Int Arch Occup Environ Health [Internet]. 2017 Jul [cited 2023 Aug 15];90(5). Available from: https://pubmed.ncbi.nlm.nih.gov/28224291/

19. Bell AC, Richards J, Zakrzewski-Fruer JK, Smith LR, Bailey DP. Sedentary Behaviour—A Target for the Prevention and Management of Cardiovascular Disease. Int J Environ Res Public Health. 2022 Dec 28;20(1):532.

20. Taylor WC, Paxton RJ, Shegog R, Coan SP, Dubin A, Page TF, et al. Impact of Booster Breaks and Computer Prompts on Physical Activity and Sedentary Behavior Among Desk-Based Workers: A Cluster-Randomized Controlled Trial. Prev Chronic Dis. 2016 Nov 17;13:E155.

21. Taylor WC, King KE, Shegog R, Paxton RJ, Evans-Hudnall GL, Rempel DM, et al. Booster Breaks in the workplace: participants’ perspectives on health-promoting work breaks. Health Educ Res. 2013 Jun;28(3):414–25.

22. Taylor WC, Williams JR, Harris LE, Shegog R. Computer Prompt Software to Reduce Sedentary Behavior and Promote Physical Activity Among Desk-Based Workers: A Systematic Review. Hum Factors. 2021 Aug 15;187208211034271.

23. Stephenson A, McDonough SM, Murphy MH, Nugent CD, Mair JL. Using computer, mobile and wearable technology enhanced interventions to reduce sedentary behaviour: a systematic review and meta-analysis. Int J Behav Nutr Phys Act. 2017 Aug 11;14(1):105.

24. Riquelme R, Rezende LFM, Marques A, Drenowatz C, Ferrari G. Association between 24-h movement guidelines and cardiometabolic health in Chilean adults. Sci Rep. 2022 Apr 6;12(1):5805.

25. Drozdz D, Alvarez-Pitti J, Wójcik M, Borghi C, Gabbianelli R, Mazur A, et al. Obesity and Cardiometabolic Risk Factors: From Childhood to Adulthood. Nutrients. 2021 Nov 22;13(11):4176.

26. Rakhmat II, Putra ICS, Wibowo A, Henrina J, Nugraha GI, Ghozali M, et al. Cardiometabolic risk factors in adults with normal weight obesity: A systematic review and meta-analysis. Clin Obes. 2022 Aug;12(4):e12523.

27. Pedisic Z, Dumuid D, Olds T. Integrating sleep, sedentary behaviour, and physical activity research in the emerging field of time-use epidemiology: Definitions, concepts, statistical methods, theoretical framework, and future directions. Kinesiology. 2017 Sep 15;49.

28. Chan AW, Tetzlaff JM, Gøtzsche PC, Altman DG, Mann H, Berlin JA, et al. SPIRIT 2013 explanation and elaboration: guidance for protocols of clinical trials. BMJ. 2013 Jan 9;346:e7586.

29. Mattsson S, Leide-Svegborn S, Andersson M. X-RAY AND MOLECULAR IMAGING DURING PREGNANCY AND BREASTFEEDING-WHEN SHOULD WE BE WORRIED? Radiat Prot Dosimetry. 2021 Oct 12;195(3–4):339–48.

30. Hurkmans CW, Knegjens JL, Oei BS, Maas AJJ, Uiterwaal GJ, van der Borden AJ, et al. Management of radiation oncology patients with a pacemaker or ICD: a new comprehensive practical guideline in The Netherlands. Dutch Society of Radiotherapy and Oncology (NVRO). Radiat Oncol Lond Engl. 2012 Nov 24;7:198.

31. López-Bueno R, Andersen LL, Koyanagi A, Núñez-Cortés R, Calatayud J, Casaña J, et al. Thresholds of handgrip strength for all-cause, cancer, and cardiovascular mortality: A systematic review with dose-response meta-analysis. Ageing Res Rev. 2022 Dec;82:101778.

32. Schokman A, Bin YS, Simonelli G, Pye J, Morris R, Sumathipala A, et al. Agreement between subjective and objective measures of sleep duration in a low-middle income country setting. Sleep Health. 2018 Dec;4(6):543–50.

33. Chase JD, Busa MA, Staudenmayer JW, Sirard JR. Sleep Measurement Using Wrist-Worn Accelerometer Data Compared with Polysomnography. Sensors. 2022 Jul 4;22(13):5041.

34. Migueles JH, Cadenas-Sanchez C, Ekelund U, Delisle Nyström C, Mora-Gonzalez J, Löf M, et al. Accelerometer Data Collection and Processing Criteria to Assess Physical Activity and Other Outcomes: A Systematic Review and Practical Considerations. Sports Med Auckl NZ. 2017 Sep;47(9):1821–45.

35. Nooijen CFJ, Blom V, Ekblom Ö, Ekblom MM, Kallings LV. Improving office workers’ mental health and cognition: a 3-arm cluster randomized controlled trial targeting physical activity and sedentary behavior in multi-component interventions. BMC Public Health. 2019 Mar 5;19:266.

36. Hees VT van, Migueles JH, Sabia S, Patterson MR, Fang Z, Heywood J, et al. GGIR: Raw Accelerometer Data Analysis [Internet]. 2023 [cited 2023 Sep 11]. Available from: https://cran.r-project.org/web/packages/GGIR/index.html

37. Bull FC, Maslin TS, Armstrong T. Global physical activity questionnaire (GPAQ): nine country reliability and validity study. J Phys Act Health. 2009 Nov;6(6):790–804.

38. Moreno-Llamas A, García-Mayor J, De la Cruz-Sánchez E. Concurrent and Convergent Validity of a Single, Brief Question for Physical Activity Assessment. Int J Environ Res Public Health. 2020 Mar;17(6):1989.

39. Chau JY, Van Der Ploeg HP, Dunn S, Kurko J, Bauman AE. Validity of the occupational sitting and physical activity questionnaire. Med Sci Sports Exerc. 2012 Jan;44(1):118–25.

40. Kastelic K, Šarabon N, Burnard MD, Pedišić Ž. Validity and Reliability of the Daily Activity Behaviours Questionnaire (DABQ) for Assessment of Time Spent in Sleep, Sedentary Behaviour, and Physical Activity. Int J Environ Res Public Health. 2022 Apr 28;19(9):5362.

41. Martin A, Fitzsimons C, Jepson R, Saunders DH, van der Ploeg HP, Teixeira PJ, et al. Interventions with potential to reduce sedentary time in adults: systematic review and meta-analysis. Br J Sports Med. 2015 Aug;49(16):1056–63.

42. Buckingham SA, Williams AJ, Morrissey K, Price L, Harrison J. Mobile health interventions to promote physical activity and reduce sedentary behaviour in the workplace: A systematic review. Digit Health. 2019 Mar 27;5:2055207619839883.

43. Cohen S, Kamarck T, Mermelstein R. A global measure of perceived stress. J Health Soc Behav. 1983 Dec;24(4):385–96.

44. Marín E, Tomás US, de Chile S. ESTANDARIZACIÓN DE LAS ESCALAS, PERCEIVED STRESS SCALE DE COHEN ET. AL. [PSS-14]. ESCALA MAGALLANES DE ESTRÉS [EMEST], Y ESTUDIO COMPARATIVO DEL RESULTADO DE AMBAS ESCALAS EN UNA MUESTRA DE PROFESIONALES DEL ÁREA METROPOLITANA DE SANTIAGO DE CHILE. 2004;

45. Kuorinka I, Jonsson B, Kilbom A, Vinterberg H, Biering-Sørensen F, Andersson G, et al. Standardised Nordic questionnaires for the analysis of musculoskeletal symptoms. Appl Ergon. 1987 Sep;18(3):233–7.

46. Prince SA, Rasmussen CL, Biswas A, Holtermann A, Aulakh T, Merucci K, et al. The effect of leisure time physical activity and sedentary behaviour on the health of workers with different occupational physical activity demands: a systematic review. Int J Behav Nutr Phys Act. 2021 Jul 20;18(1):100.

47. Pernambuco CS, Laranjeira C, Mesquita MG, Conceição M da, Souza VRG de, Dantas EM. CONCENTRACION URINARIA DE HIDROXIPROLINA EN HOMBRES CON LUMBALGIA SOMETIDOS A HIDROKINESIOTERAPIA. J Mov Health [Internet]. 2010 Aug 31 [cited 2023 Aug 15];11(2). Available from: http://jmh.pucv.cl/index.php/jmh/article/view/28

48. Khan MS, Aouad R. The Effects of Insomnia and Sleep Loss on Cardiovascular Disease. Sleep Med Clin. 2022 Jun;17(2):193–203.

49. Martínez-González MA, García-Arellano A, Toledo E, Salas-Salvadó J, Buil-Cosiales P, Corella D, et al. A 14-Item Mediterranean Diet Assessment Tool and Obesity Indexes among High-Risk Subjects: The PREDIMED Trial. PLoS ONE. 2012 Aug 14;7(8):e43134.

50. Barnard ND, Alwarith J, Rembert E, Brandon L, Nguyen M, Goergen A, et al. A Mediterranean Diet and Low-Fat Vegan Diet to Improve Body Weight and Cardiometabolic Risk Factors: A Randomized, Cross-over Trial. J Am Nutr Assoc. 2022 Feb;41(2):127–39.

51. Heatherton TF, Kozlowski LT, Frecker RC, Fagerström KO. The Fagerström Test for Nicotine Dependence: a revision of the Fagerström Tolerance Questionnaire. Br J Addict. 1991 Sep;86(9):1119–27.

52. Bernal NG. Distribución e ingreso autónomo promedio de hogares según decil de ingreso. 2021;

53. Clawson AH, Nwankwo CN, Baraldi AN, Cole AB, Berlin KS, Ruppe NM, et al. Longitudinal smoking patterns and adult cardiometabolic risk among African Americans. Health Psychol Off J Div Health Psychol Am Psychol Assoc. 2021 Jan;40(1):51–61.

54. Kempel MK, Winding TN, Böttcher M, Andersen JH. Subjective social status and cardiometabolic risk markers in young adults. Psychoneuroendocrinology. 2022 Mar;137:105666.

55. Asoción Chilena de Seguridad. Ejercicios para descansar cuello, hombros y vista. 2022;

56. Asoción Chilena de Seguridad. Ejercicios para descansar brazos y piernas. 2022;

57. Scherr J, Wolfarth B, Christle JW, Pressler A, Wagenpfeil S, Halle M. Associations between Borg’s rating of perceived exertion and physiological measures of exercise intensity. Eur J Appl Physiol. 2013 Jan;113(1):147–55.

58. Turner EL, Li F, Gallis JA, Prague M, Murray DM. Review of Recent Methodological Developments in Group-Randomized Trials: Part 1-Design. Am J Public Health. 2017 Jun;107(6):907–15.

59. Rutterford C, Copas A, Eldridge S. Methods for sample size determination in cluster randomized trials. Int J Epidemiol. 2015 Jun;44(3):1051–67.

60. Kohl HW, Craig CL, Lambert EV, Inoue S, Alkandari JR, Leetongin G, et al. The pandemic of physical inactivity: global action for public health. Lancet Lond Engl. 2012 Jul 21;380(9838):294–305.

61. Lally P, van Jaarsveld CHM, Potts HWW, Wardle J. How are habits formed: Modelling habit formation in the real world. Eur J Soc Psychol. 2010;40(6):998–1009.

